# A Network-Based Stochastic Epidemic Simulator: Controlling COVID-19 with Region-Specific Policies

**DOI:** 10.1101/2020.05.02.20089136

**Authors:** Askat Kuzdeuov, Daulet Baimukashev, Aknur Karabay, Bauyrzhan Ibragimov, Almas Mirzakhmetov, Mukhamet Nurpeiissov, Michael Lewis, Huseyin Atakan Varol

## Abstract

In this work, we present an open-source stochastic epidemic simulator, calibrated with extant epidemic experience of COVID-19. Our simulator incorporates information ranging from population demographics and mobility data to health care resource capacity, by region, with interactive controls of system variables to allow dynamic and interactive modeling of events. The simulator can be generalized to model the propagation of any disease, in any territory, but for this experiment was customized to model the spread of COVID-19 in the Republic of Kazakhstan, and estimate outcomes of policy options to inform deliberations on governmental interdiction policies.

## I. Introduction

CORONAVIRUS disease 2019 (COVID-19) has emerged as a global crisis that threatens to overwhelm public healthcare systems and disrupt the social and economic welfare of every country. The daily lives of billions of people have been impacted by the unprecedented social controls imposed by governments to inhibit the spread of the novel coronavirus [1]-[3].

COVID-19 has spread rapidly amongst a globally susceptible population, with rates of propagation and mortality greater than the averages typically associated with influenza [4]. Initial projections of the global impact [5] indicate that it will likely become the most severe pandemic event over a century, dating to the influenza epidemic of 1918.

The situation is exacerbated by several unfortunate observations: the population has no prior exposure, carriers are highly contagious in pre-symptomatic states [6], there is no vaccine yet available, widespread testing for the virus began quite late due to lack of testing kits, reagents, and facilities, and there appears to be broad variation of risk profiles based on population demographics and prior health history [7].

In this scenario, epidemiological models can be used to project the future course of the disease, and to estimate the impact of non-pharmaceutical interventions (NPIs) and related control measures that might be used to slow the contagion, and thereby provide time to enhance health care resources and develop effective immunological defenses such as new vaccines.

We have developed and implemented a network-based stochastic epidemic simulator (leveraging our prior work [8]) which models cities and regions as nodes in a graph, and the edges between nodes representing transit links of roads, railways, and air travel routes to model the mobility of inhabitants amongst cities. The simulator includes population demographics along with health care system capacity, in particular, the intensive care unit (ICU) availability, which serves as a negative impact multiplier when the number of ICU beds is exceeded.

In each node, the simulator runs a compartmental Susceptible-Exposed-Infectious-Recovered (SEIR) model, such that individuals can cycle through the four stages based on state transition probabilities. These probabilities are based on parameters such as the susceptible-to-exposed transition constant and the mortality rate, which can be influenced by age, gender, genetic profile, and health status.

The simulator can be used to estimate the extent and duration of an epidemic over time, and model the potential impact of NPI measures deployed to suppress or mitigate the spread of the virus. The supported NPI control measures include what are commonly described as social (or physical) distancing, such as limiting travel or quarantining a region, on a localized basis and by transit link.

## II. Related Work

The Susceptible-Infected-Recovered (SIR) epidemic model is one of the first proposed for epidemiological simulation purposes, and has been utilized widely due to its simplicity and effectiveness [9]. In the SIR model, a society consists of three compartments. The first compartment, susceptible (S), contains individuals who are vulnerable and not yet infected. The second compartment, infected (I), is formed from susceptible individuals who become infected; in this state, they are capable of shedding the virus and spreading the disease. The last compartment, recovered (R) consists of previously infected individuals who have overcome the disease. The recovered individuals are presumed to have acquired some level of immunity to the disease, such that they have a lower probability of reinfection compared to susceptible individuals.

The SIR model, however, has several limitations based on simplifying assumptions that do not correlate well to actual viral propagation. For instance, the SIR model assumes that each individual has an equal probability of spreading the disease, and that infected individuals themselves become infectious immediately, when, in practice, there is often a latency period in between. The model also presumes that each region has a fixed-size population, thus not accounting for population mobility.

The experience with the SIR model has motivated many enhancements and variations, with new compartmental states, such as SEIR, SEIRS, SIRS, SEI, SEIS, SI, and SIS [10]. Many of these models incorporate the additional compartment - exposed (E). In one variation, individuals are infected in this Exposed state but not yet infectious during the latent period; these individuals become infectious when the latent period ends. In general, compartments and the related state transition probabilities are selected based on the characteristics of the specific disease and the purpose of the model.

The basic reproduction rate *R*_0_, denoting the average number of individuals infected by an infectious individual, is used as a parameter in many epidemiology models [10]. It can be interpreted as the ability of the infection to intrude and persist in a new population. In deterministic models, infection spreads in a sustained manner among a susceptible population if *R*_0_ *>* 1 [11].

The propagation of infectious diseases can be simulated using both deterministic and stochastic models [12]. A deterministic epidemic model is formulated as a system of differential equations, while a stochastic epidemic model can be implemented using stochastic differential equations or discrete time Markov processes [13]. The deterministic model yields the same solution each time if the initial conditions and parameter values of the model are not changed. The distinction of a stochastic model is that it contains at least one probabilistic element, i.e. the output of the model varies at some range even if the initial conditions and parameter values of the model remain constant.

Both approaches have their advantages and limitations [14], [15]. The main strength of the deterministic model is that it clearly shows how the initial conditions and parameters impact the model behavior, but at the expense of more realistic and nuanced simulation. A deterministic model ignores uncertainties, which can be considered as a major limitation due to the fact that epidemic propagation is inherently a stochastic phenomenon. In contrast, the stochastic models take into account uncertainties, though by introducing probabilities, with the natural outcome that no two simulations produce exactly the same results, stochastic models are regarded as more challenging to manage and demonstrate causal links.

Variations of deterministic and stochastic models, in the context of both fixed-population and networked dynamics, have been studied in the literature [16]. A stochastic ISIR epidemic model with immunization strategies was presented to simulate the epidemic spread on social contact networks [17]. The SEIR model with time delay on scale-free networks was described in [18] to investigate the reproduction number and transmission dynamics of the disease. A model that analyses a population with two diseases, infectious and noninfectious, was introduced in [19]. The model assumes that the non-infectious disease itself is not dangerous but capable to attack the immune system, which means that its combination with infectious diseases might be fatal.

In our work, the main focus is to develop a network-based simulator that closely models the population and regional mobility dynamics. Therefore, we employed the SEIR stochastic model to simulate the spread of COVID-19 virus in a closed population. Then, we extended the model to investigate the dynamics of the spread of the disease between different regions. The detailed methodology is provided in Section III.

In order to make epidemic simulators more realistic, it is important to populate them with authentic initial conditions, and calibrate the system variables against real data. Prior examples include the SEIR model that was implemented using the Anylogic program to examine the spread of H1N1 influenza virus between different regions of Korea based on the highway and domestic flight traffic data [20]. In [21], the SEIR model was introduced which takes into account a number of workers in a region, their geographic locations and age groups. An agent-based system that uses social interactions and individual mobility patterns was employed to study the H1N1 outbreak in Mexico [22].

For our purposes, to accurately model the situation of the Republic of Kazakhstan as our case study, we incorporated transit data from highway travel, domestic flights, and railway connections amongst the regions of the country. In addition, we used the actual population demographics and hospital capacities of each region, provided under a collaboration agreement with the government.

One of the most important stages in the development of epidemiological models is the calibration of system variables. Calibration is tuning model parameters with available real epidemic data. This process is necessary to refine the model’s ability to accurately project future scenarios in a reasonable and demonstrable manner, which can then be compared with actual events in real time to either affirm or undermine the model’s assumptions. With sufficient data, particle and ensemble filters can be utilized to further improve model accuracy [23], [24]. In our case, COVID-19 is at an early stage of spreading in Kazakhstan. The first official case was announced on the 13 March 2020 [25]. Therefore, we calibrated our model empirically, first with reference to prior examples, and then with the limited available case data for the country.

Another important feature of epidemiological simulation is the visualization of the outcomes. The visualizations help users better understand the propagation of the virus, and the impact of policy decisions. There are many publicly available software tools for epidemic simulation with graphical interface components [26], [27].

Many countries have developed their own COVID-19 epidemic models to simulate different scenarios [28]–[32]. The main advantage of our epidemic simulator is that the source code is publicly available, thus researchers can use or adapt it to their specific problem and save valuable time. Also, we provide a visualization tool such that users without programming skills can easily run the simulator and readily comprehend the results. The graphical user interface allows the user to set the initial conditions and model parameters, to dynamically adjust parameters during model execution, to incrementally plot the simulation results in real-time, to save the intermediate and final results, and to upload the saved model and continue the simulation. In addition, we have prepared a series of video tutorials^1^ providing background context for the project, describing the architecture of the model, the implementation and use of the simulator, and the outcomes from the assessment of the Republic of Kazakhstan scenarios.

## III. Methodology

### A. Stochastic Epidemic Simulator for Single Nodes

In our simulator, the SEIR model for a single node consists of four superstates (Susceptible (*S^S^*), Exposed (*E^S^*), Infected (*I^S^*) and Recovered (*R^S^*)) and transitions amongst states occur in accordance with the statechart shown in Fig. 1. A description of parameters is given in the Table I. The Susceptible superstate (*S^S^*) consists of two states: Susceptible (*S*) and Vaccinated (*V*). The daily vaccination rate *vr* dictates the transition between these states. The ratio of immunized people after vaccination *vir* is used to describe the portion of individuals going from Vaccinated state (*V*) to Vaccination Immunized (*VI*) or back to the Susceptible (*S*); however, it is not yet activated since there is no known vaccine for COVID-19. In order to represent delay dynamics of epidemics, exposure *t_exp_*, infection *t_inf_*, and vaccination immunization tvac periods are taken into account. The expected value (*EV*) of the transition from Susceptible state (*S*) to Exposed state (*E*) is accomplished according to the parameter β*_exp_*, as

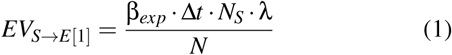

where ∆*t* is a simulation step size, *N_S_* is the total number of individuals in Susceptible state (*S*), *N* is the total population, and λ is a weighted population which is estimated as

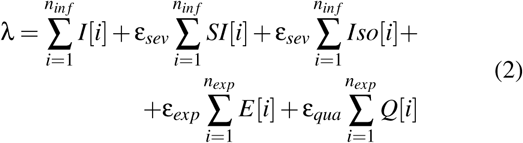

where *n_exp_* and *n_inf_* are the total number of substates in the Exposed (*E*) and Infected (*I*) superstates, and *Iso* is Isolated state, *SI* is Severe Infected state, and *Q* is Quarantined state. Along with the total number of substates of the Vaccination state *n_vac_*, these are calculated as

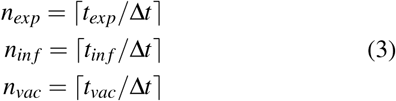

**Fig. 1:**
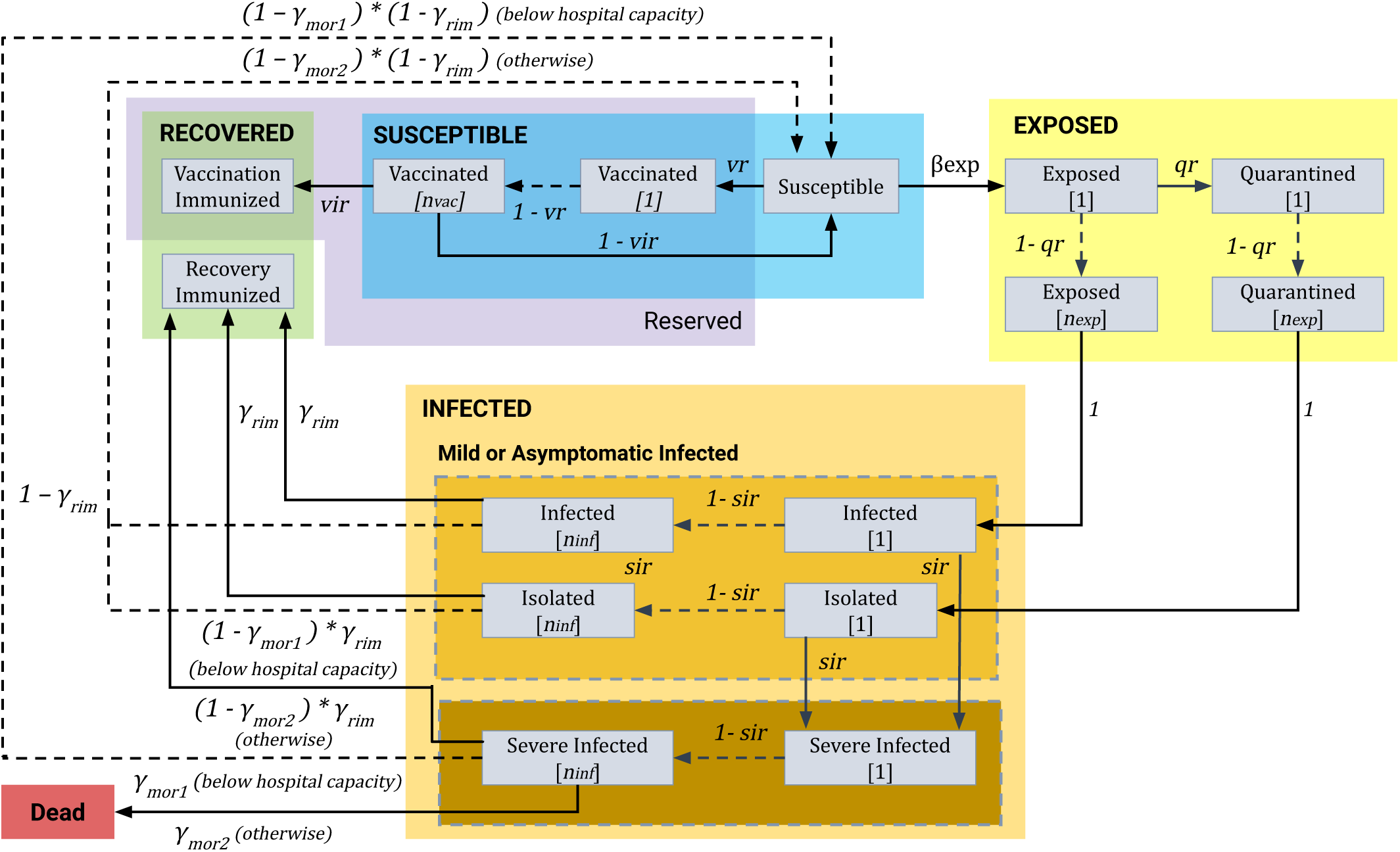
The statechart of the SEIR epidemic node simulator.

**Fig. 2:**
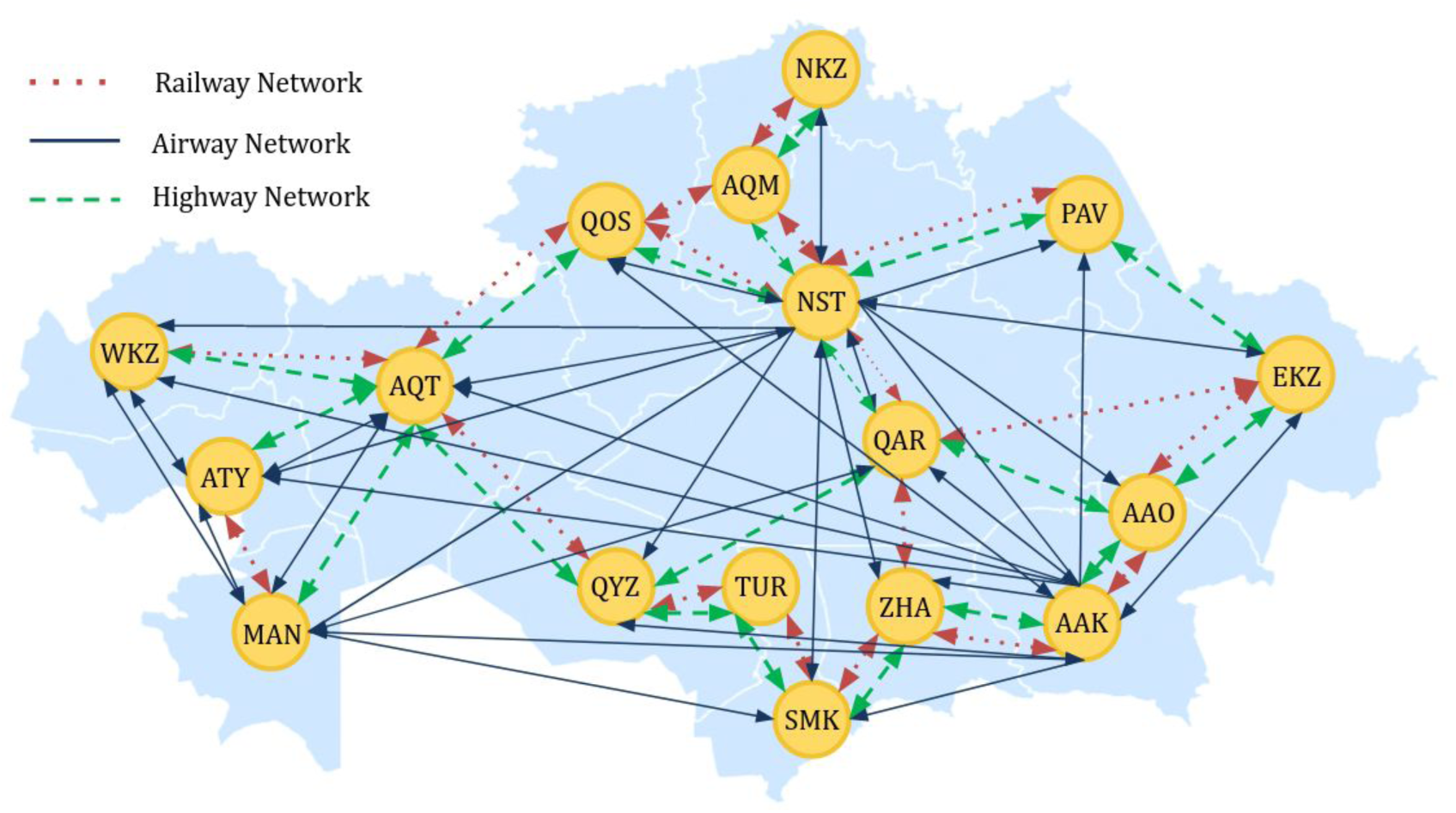
Kazakhstan as a network: Nodes representing the administrative divisions (regions and big cities) of the country are overlaid on the map. The edges of the network represent the highway, railway, and airway connections between different administrative divisions.

**Table I:** List of simulation parameters and their descriptions.

| Parameter | Description |
| --- | --- |
| $vr$ | Daily vaccination rate |
| $vir$ | Vaccination immunization rate |
| $\beta_{exp}$ | Susceptible to Exposed transition constant |
| $qr$ | Daily quarantine rate (Ratio of Exposed individuals getting Quarantined) |
| $sir$ | Daily severe infection rate (Ratio of Infected or Isolated individuals getting Severe Infected) |
| $hc$ | Hospital capacity |
| $\gamma_{rim}$ | Infected to Recovery Immunized transition constant |
| $\gamma_{mor1}$ | Severe Infected to Dead transition probability (if the hospital capacity is not exceeded) |
| $\gamma_{mor2}$ | Severe Infected to Dead transition probability (if the hospital capacity is exceeded) |
| $\epsilon_{exp}$ | Transmission rate of Exposed compared to the Infected |
| $\epsilon_{qua}$ | Transmission rate of Quarantined compared to the Infected |
| $\epsilon_{sev}$ | Transmission rate of Severe Infected compared to the Infected |
| $t_{exp}$ | Exposure period in days |
| $t_{inf}$ | Infection period in days |
| $t_{vac}$ | Vaccination immunization period in days |
| $\Delta t$ | Sampling time, in days, for updating states |

The Exposed superstate (*E^S^*) includes Exposed (*E*) and Quarantined (*Q*) states and the transition between these states is governed by the daily quarantine rate *qr*. The transition rates between the Exposed superstate (*E^S^*) and Infected superstate (*I^s^*) are equal to one since after the end of incubation period all individuals transfer from Exposed (*S*) and Quarantine (*Q*) states to Infected (*I*) and Isolated (*Iso*) states respectively.

The main difference from our previous model, presented in [8], is the introduction of a third state Severe Infected (*SI*) within the Infected superstate (*I^s^*). Individuals move from the Infected (*I*) and Isolated (*Iso*) states to the Severe Infected (*SI*) according to the severe infection rate *sir*.

The natural death rate is neglected in Infected (*I*) and Isolated (*Iso*) states and the transition to the Dead state (*D*) occurs only through the Severe Infected state (*SI*). This is done to model the high number of asymptomatic or mild cases which carry the disease. The natural birth and death rate factors are removed due to their negligible impact over the relatively short duration of the simulation. Also, individuals move from Infected (*I*) and Isolated (*Iso*) states to Recovery Immunized (*RI*) state according to the recovery immunization rate *γ_rim_*, and the rest make a transition to the Susceptible state (*S*).

The mortality rate used in the model is divided into γ*_mor_*_1_ and γ*_mor_*_2_ depending on the hospital capacity: γ*_mor_*_1_ is applied when the number of people in Severe Infected state (*SI*) is below the hospital capacity, otherwise the model uses γ*_mor2_* mortality rate which is greater than γ_mor1_. Finally, the individuals move from Severe Infected state (*SI*) to Recovery Immunized (*RI*) and Susceptible (*S*) states. In case, if the hospital capacity is not exceeded then the expected transition values are calculated as

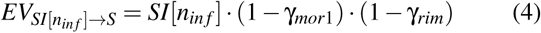

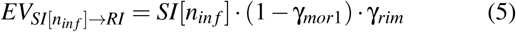

otherwise,

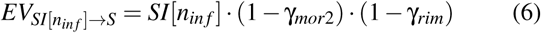

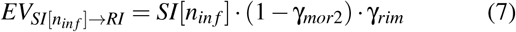

In order to launch a run of the simulator, the values of transition parameters and the number of individuals in each state must be initialized, after which the stochastic solver will proceed with the iterations of time units, generating the corresponding population states based on those parameters.

### B. A Network Model for Epidemic Simulation

A network model in the epidemiological context is the representation of a country as a graph of interconnected nodes, where an individual node represents an administrative unit of the country, such as a city or region, taking into account their respective population demographics and health care capacities. This is a convenient representation since measures during epidemics are usually taken for specific administrative units. The transportation connections between the regions can also be modeled as the edges of the network. Each node in the network runs concurrently as a separate instance of the single-node SEIR model described in Sec. III-A. The internal parameters of each SEIR model can be changed independently. The network model thus allows for viral propagation, with corresponding state transitions, within and amongst the nodes representing the cities and the regions. For a network with *M* nodes, the total transition matrix 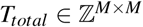 can be created by collecting and combining the open source data on the daily domestic air (*T_air_*), rail (*T_rail_*), and highway (*T_highway_*) travel between different regions as

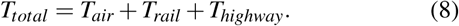

To simulate the network transition between nodes, at each sampling time of the network simulator ∆*T*, the randomly sampled population is transferred from one node to another according to the transition matrix. It is important to note that the sampling times ∆*T* used for the network and ∆*t* for the node are different (∆*T* > ∆*t*). The expected value of transition from state *K* of node *i* to the corresponding state *K* of node *j* is given by the following equation:

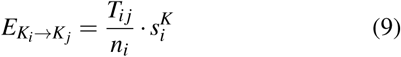

where *T_ij_* is the population transfer from node *i* to node *j*, *n_i_* is the population of node *i*, 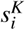 is the number of individuals in the state *K* of node *i*. Some of the states such as Quarantined and Severe Infected are excluded from the transfer between the nodes, i.e. *K* ∉ *{Q,SI,Iso,D}*.

Our simulator allows disconnection of a particular node by turning it off from the list of air, highway and railway transitions. Also, the traffic *tr* and leakage ratios *lr* facilitate the fine tuning of the transportation between the nodes. Specifically, by lowering the traffic ratio, the number of individuals transferring between the nodes can be decreased. This can be used to simulate the policies which throttle down the transportation between regions. The leakage ratio allows minor transitions even though a node is disconnected from the network. This is done to take into account individuals that cross quarantined borders illegally and also for transportation between regions for essential services such as food delivery.

### C. Simulator Implementation

The simulator was implemented in Python programming language. For faster operation, we used the *multiprocessing* module of Python to parallelize the execution of the single node simulations across all regions. A graphical user interface (GUI) for the simulator (see Fig. 3) was developed using the interactive visualization library *Bokeh* [33]. This open-source BSD license library utilizes web-browsers to visualize data. This simplifies online deployment of the simulator and also enables platform independence. The source code^2^ of our simulator was uploaded to GitHub under the BSD license.

**Fig. 3:**
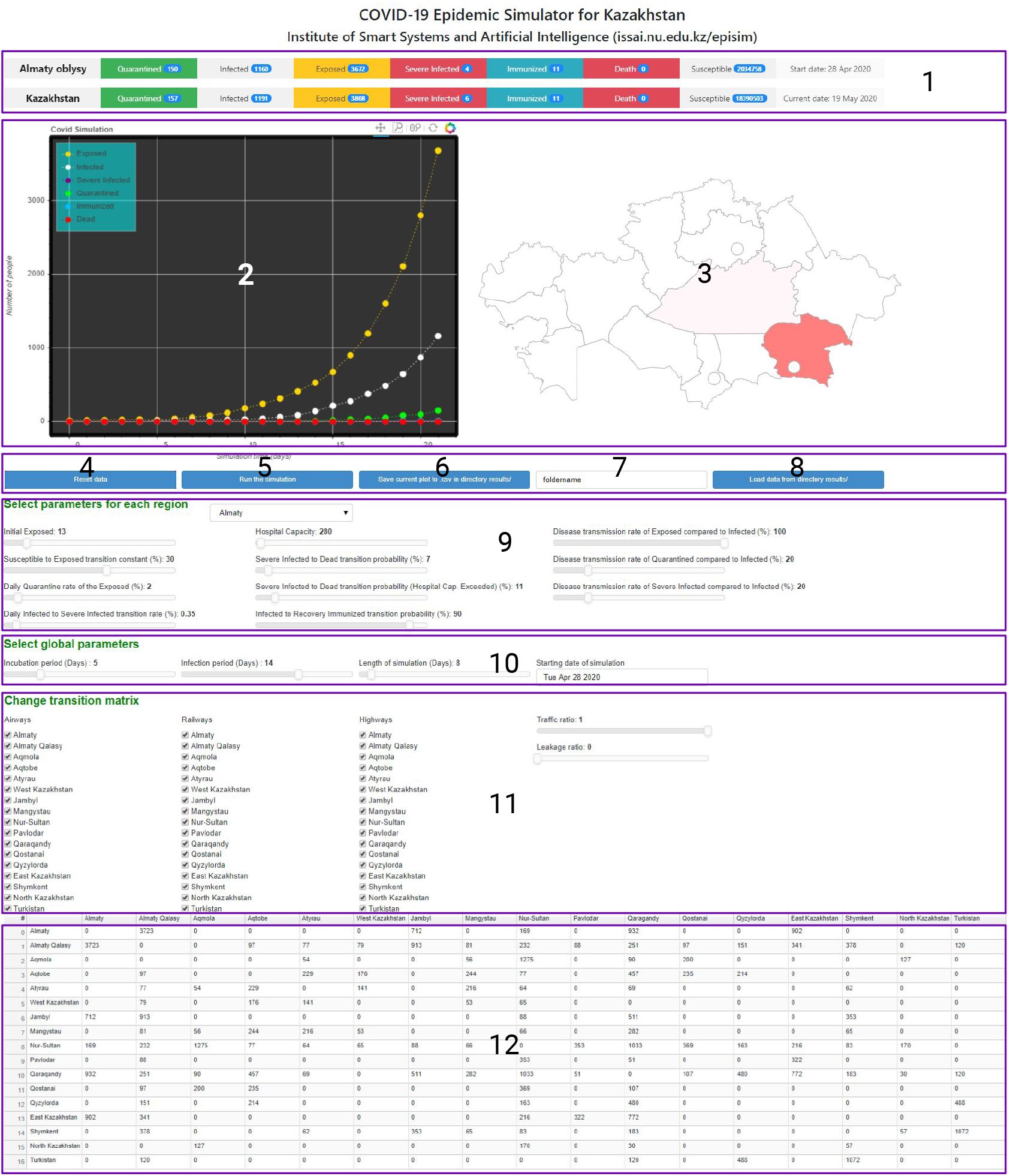
The graphical user interface (GUI) of the simulator: 1 - Current values of states for a selected region and the whole country, 2 - Plot showing the time evolution of the states for a selected node, 3 - Interactive map of the country (in this case Republic of Kazakhstan), 4 - Button to reset current parameters, 5 - Button to start the simulation, 6 - Button for saving results and model parameters, 7 - Text box for inserting the path to save files, 8 - Button for loading a saved simulation, 9 - Sliders to set the parameters of a selected region, 10 - Sliders for choosing the global simulation parameters, 11 - Checkboxes to configure the transition matrix, and 12 - Transition matrix.

Figure 3 illustrates the components of the simulator interface. The number of individuals in each state for a selected region and for the whole country are shown on the top side (1) in real-time. Also, the starting date of the simulation and the current date are provided here. The epidemic dynamics versus time are visualized in the upper left plot (2). On the right side of this plot, the epidemic heat map (3) is displayed. Also, the GUI allows to save simulation results and model parameters to a *csv* file (6). The saved model parameters can be reloaded (7, 8) to continue the simulation from a prior point. Also, it is possible to reset the parameters to default values (4). Simulation parameters can be selected for each node (9) and for the whole network (10). The transition matrix can be adjusted using checkboxes (11) and slider bars to adjust traffic and leakage ratios. The resulting transition matrix is shown at the bottom part of the interface (12).

## IV. Simulations and Results

### A. Single Node Epidemic Simulation of Lombardy (Italy)

In order to test the capability of our simulator and fine tune its parameters, we first focused on the simulation of the Lombardy region in Italy. We chose Lombardy since it is the epicenter of the COVID-19 outbreak in Italy, its epidemic timeline is well-established, and the Italian government has shared comprehensive daily epidemic data since 24 February 2020 [34], thus facilitating the calibration of model assumptions against actual events and data. The population of Lombardy is just over 10 million people, therefore, we used this number to initialize the number of susceptible individuals in our model. We started the simulation on 1 January 2020 based on the results presented in [35]. We set the number of exposed individuals to ten on this date, as this figure yielded accurate model state populations going forward. In order to tune other parameters of the model, we used the reported data for the Lombardy region for the period between 24 February and 27 April 2020. Also, we took into account the timeline of events and government policy decisions that would impact the simulation (see Table II).

**Table II:** Lombardy COVID-19 Timeline

| Day | Date | Death | Confirmed cases | Event |
| --- | --- | --- | --- | --- |
| 0 | 1/1/2020 | unk. | 1 | The first confirmed case [35]. |
| 54 | 24/2/2020 | 6 | 172 | The COVID-19 data repository was launched [34]. |
| 61 | 2/3/2020 | 38 | 1,254 | "Red zone" in Lombardy. |
| 67 | 8/3/2020 | 267 | 4,189 | Whole Lombardy locked down. |
| 70 | 11/3/2020 | 617 | 7,280 | Bars, restaurants are closed. |
| 81 | 22/3/2020 | 3,456 | 27,206 | Factories and all nonessential productions are closed. |
| 118 | 27/4/2020 | 13,449 | 73,479 | Date of last data available. |

The model parameters are summarized in Table III. A parameter β*_exp_* was tuned according to the events described in Table II. The model was simulated for 180 days with the sampling time ∆*t* = 1/24 days.

The results of the simulation are shown in Fig. 4. It can be seen from the figure that the model fits the reported number of deaths for the given period of time closely. Also, we can observe that the epidemic reaches a peak around 26 March 2020 when the total number of infected individuals is 67,850. If we look at the reported data, new daily confirmed cases increased significantly starting from 19 March 2020 (2171 cases) and remained high until 28 March 2020 (2117 cases). After this period, the new daily cases started to gradually decrease. Thus, we can assume that the model estimated the peak of the spread of COVID-19 accurately. Also, according to the simulation, we forecast that the epidemic might continue until the end of June and the number of total deaths might exceed 17,000 in Lombardy.

**Table III:**
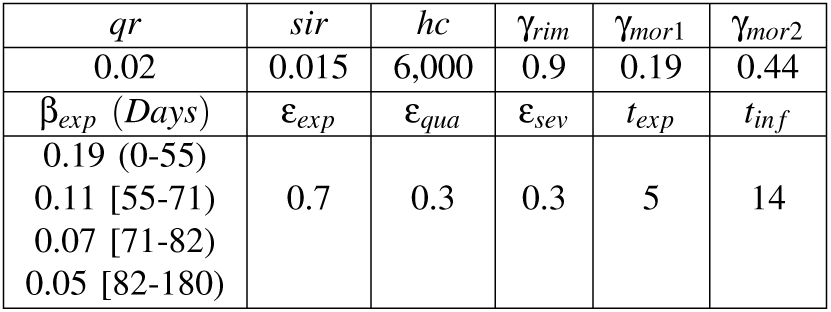
Single Node Simulation Parameters for Lombardy.

**Fig. 4:**
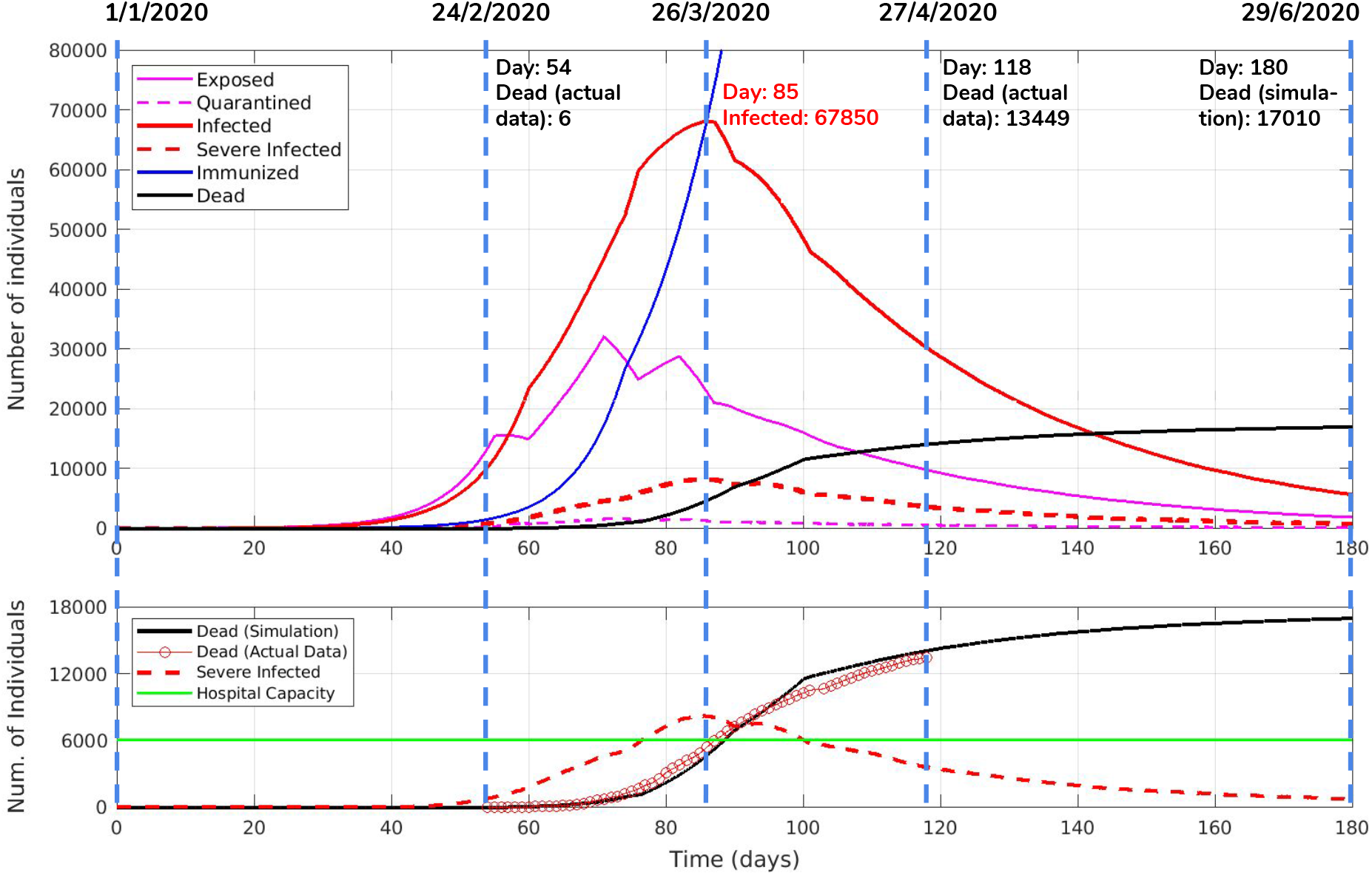
Simulation results for the Lombardy region. Upper plot shows the states of the epidemic simulation versus time. The vertical dashed lines indicate events from the timeline in the Table II. Bottom plot shows the number of Severe Infected individuals along with the hospital capacity and also compares the number of deaths in the simulation to the actual number of deaths due to COVID-19.

### B. Network-based Simulation of the COVID-19 in Kazakhstan

#### 1) Model Parameters and Validation

As noted, we tested the model’s assumptions against prior epidemic experience, and then customized the model to simulate the spread of COVID-19 in the Republic of Kazakhstan. We performed model calibration and validation by replicating the observed COVID-19 numbers in the country.

Epidemics usually start in hubs with the import of the disease from another country and propagates to other regions via transportation links between regions. The first confirmed instances of COVID-19 patients in Kazakhstan were detected in the largest cities, Nur-Sultan and Almaty, and then the disease spread to other regions [25]. These two cities serve as transportation hubs as highlighted in Fig. 2.

Table IV lists all nodes with the respective abbreviations, population numbers and hospital capacities for the Republic of Kazakhstan. The transition matrix between nodes representing the domestic movements in the country have been estimated based on the government records and available statistical data (see Table V), and captures the end destination numbers, not accounting for intermediate transit points. For instance, if one travels from the capital to the other regions via a third transfer city, only the departure and arrival nodes have been considered in the transition matrix.

**Table IV:** Names, abbreviations, population and hospital capacity of the nodes.

| Node | Name | Abbr. | Population | Hospital capacity |
| --- | --- | --- | --- | --- |
| 1 | Almaty | AAO | 2,039,376 | 280 |
| 2 | Almaty Qalasy | AAK | 1,854,556 | 2,395 |
| 3 | Aqmola | AQM | 738,587 | 895 |
| 4 | Aqtobe | AQT | 869,603 | 600 |
| 5 | Atyrau | ATY | 633,801 | 650 |
| 6 | West Kazakhstan | WKZ | 652,314 | 250 |
| 7 | Zhambyl | ZHA | 1,125,297 | 725 |
| 8 | Mangystau | MAN | 678,224 | 100 |
| 9 | Nur-Sultan | NST | 1,078,362 | 885 |
| 10 | Pavlodar | PAV | 753,804 | 425 |
| 11 | Qaragandy | QAR | 1,378,554 | 1,670 |
| 12 | Qostanay | QOS | 872,736 | 300 |
| 13 | Qyzylorda | QYZ | 794,165 | 465 |
| 14 | East Kazakhstan | EKZ | 1,378,504 | 1,420 |
| 15 | Shymkent | SMK | 1,011,511 | 1,505 |
| 16 | North Kazakhstan | NKZ | 554,519 | 380 |
| 17 | Turkistan | TUR | 1,981,747 | 300 |

**Table V:** The total transition matrix between the nodes which is the sum of the domestic air, highway and rail transition matrices.

| Node | AAO | AAK | AQM | AQT | ATY | WKZ | ZHA | MAN | NST | PAV | QAR | QOS | QYZ | EKZ | SMK | NKZ | TUR |
| --- | --- | --- | --- | --- | --- | --- | --- | --- | --- | --- | --- | --- | --- | --- | --- | --- | --- |
| AAO | 0 | 7,447 | 0 | 0 | 0 | 0 | 1424 | 0 | 338 | 0 | 1,865 | 0 | 0 | 1805 | 0 | 0 | 0 |
| AAK | 7,447 | 0 | 0 | 195 | 154 | 158 | 1827 | 163 | 465 | 176 | 503 | 195 | 303 | 683 | 756 | 0 | 240 |
| AQM | 0 | 0 | 0 | 0 | 109 | 0 | 0 | 113 | 2,551 | 0 | 180 | 400 | 0 | 0 | 0 | 254 | 0 |
| AQT | 0 | 195 | 0 | 0 | 459 | 352 | 0 | 448 | 154 | 0 | 914 | 471 | 429 | 0 | 0 | 0 | 0 |
| ATY | 0 | 154 | 109 | 459 | 0 | 283 | 0 | 431 | 128 | 0 | 139 | 0 | 0 | 0 | 124 | 0 | 0 |
| WKZ | 0 | 158 | 0 | 352 | 283 | 0 | 0 | 106 | 130 | 0 | 0 | 0 | 0 | 0 | 0 | 0 | 0 |
| ZHA | 1,424 | 1,827 | 0 | 0 | 0 | 0 | 0 | 0 | 176 | 0 | 1,023 | 0 | 0 | 0 | 706 | 0 | 0 |
| MAN | 0 | 163 | 113 | 488 | 431 | 106 | 0 | 0 | 133 | 0 | 565 | 0 | 0 | 0 | 130 | 0 | 0 |
| NST | 338 | 465 | 2,551 | 154 | 128 | 130 | 176 | 133 | 0 | 706 | 2067 | 738 | 326 | 433 | 167 | 340 | 0 |
| PAV | 0 | 176 | 0 | 0 | 0 | 0 | 0 | 0 | 0 | 706 | 103 | 0 | 0 | 645 | 0 | 0 | 0 |
| QAR | 1,865 | 503 | 180 | 914 | 139 | 0 | 1,023 | 565 | 2,067 | 103 | 0 | 214 | 960 | 1,545 | 366 | 60 | 240 |
| QOS | 0 | 195 | 400 | 471 | 0 | 0 | 0 | 0 | 738 | 0 | 214 | 0 | 0 | 0 | 0 | 0 | 0 |
| QYZ | 0 | 303 | 0 | 429 | 0 | 0 | 0 | 0 | 326 | 0 | 960 | 0 | 0 | 0 | 0 | 0 | 977 |
| EKZ | 1,805 | 683 | 0 | 0 | 0 | 0 | 0 | 0 | 433 | 645 | 1,545 | 0 | 0 | 0 | 0 | 0 | 0 |
| SMK | 0 | 765 | 0 | 0 | 124 | 0 | 0 | 706 | 130 | 167 | 0 | 366 | 0 | 0 | 0 | 114 | 2,144 |
| NKZ | 0 | 0 | 254 | 0 | 0 | 0 | 0 | 0 | 340 | 0 | 60 | 0 | 0 | 0 | 114 | 0 | 0 |
| TUR | 0 | 240 | 0 | 0 | 0 | 0 | 1,384 | 0 | 0 | 0 | 240 | 0 | 977 | 0 | 2,144 | 0 | 0 |

The timeline, introduced in Table VI, of the governmental policies related to COVID-19 have been used to dynamically adjust model parameters. Since the first confirmed-positive cases are dated to 13 March 2020 in Nur-Sultan and Almaty cities, we started the simulation from 1 March 2020 with 10 exposed individuals in each of these two cities. The simulation parameters are summarized in Table VII. The node and network sampling times were set as ∆*t* = 1/24 and ∆*T* = 1/2 days.

**Table VI:** Kazakhstan COVID-19 Simulation Scenario Based on the Timeline from 1 March to 27 April 2020 [25].

| Day | Date | Death | Confirmed cases | Event |
| --- | --- | --- | --- | --- |
| 0 | 1/3/2020 | 0 | 0 | Start of the simulation. |
| 12 | 13/3/2020 | 0 | 3 | The first confirmed cases. |
| 18 | 19/3/2020 | 0 | 49 | "Red zone" in NST and AAK. |
| 21 | 22/3/2020 | 0 | 56 | NST and AAK locked down. |
| 26 | 27/3/2020 | 0 | 150 | "Red zone" in 6 other regions. |
| 30 | 31/3/2020 | 2 | 340 | The whole country locked down. |
| 57 | 27/4/2020 | 25 | 2,860 | Date of last data available at the time of writing the paper. |

**Table VII:**
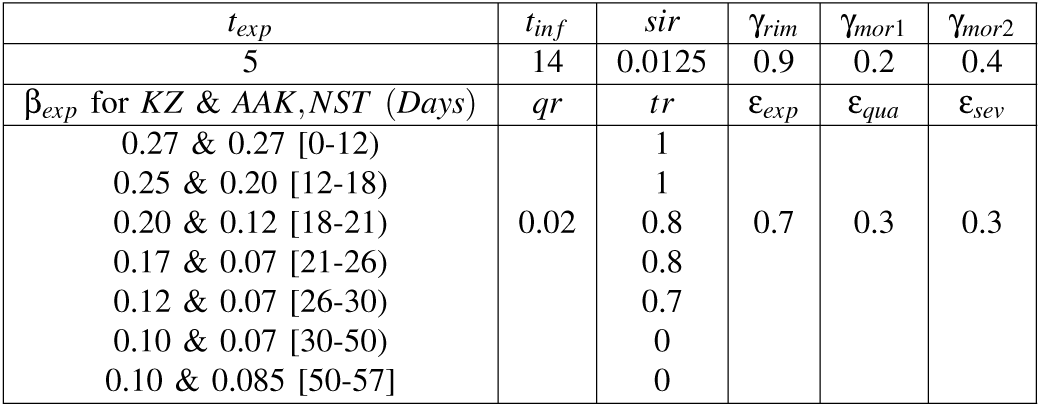
Network-based Simulation Parameters for Kazahstan.

The simulation results are shown Fig. 5. It can be seen that, on 27 April 2020, the actual number of COVID-19 deaths is 25 while the predicted number of deaths is 26. This suggests that the model was calibrated accurately and can be utilized to simulate future scenarios. As for the Almaty city result, the simulation concludes 12 dead and 1188 infected by April 27. Whereas, the reported statistics are 8 and 876 respectively. The discrepancy might be due to the stochastic nature of the simulator and the lack of testing available in the city which might result in lower number of the officially registered cases [25].

**Fig. 5:**
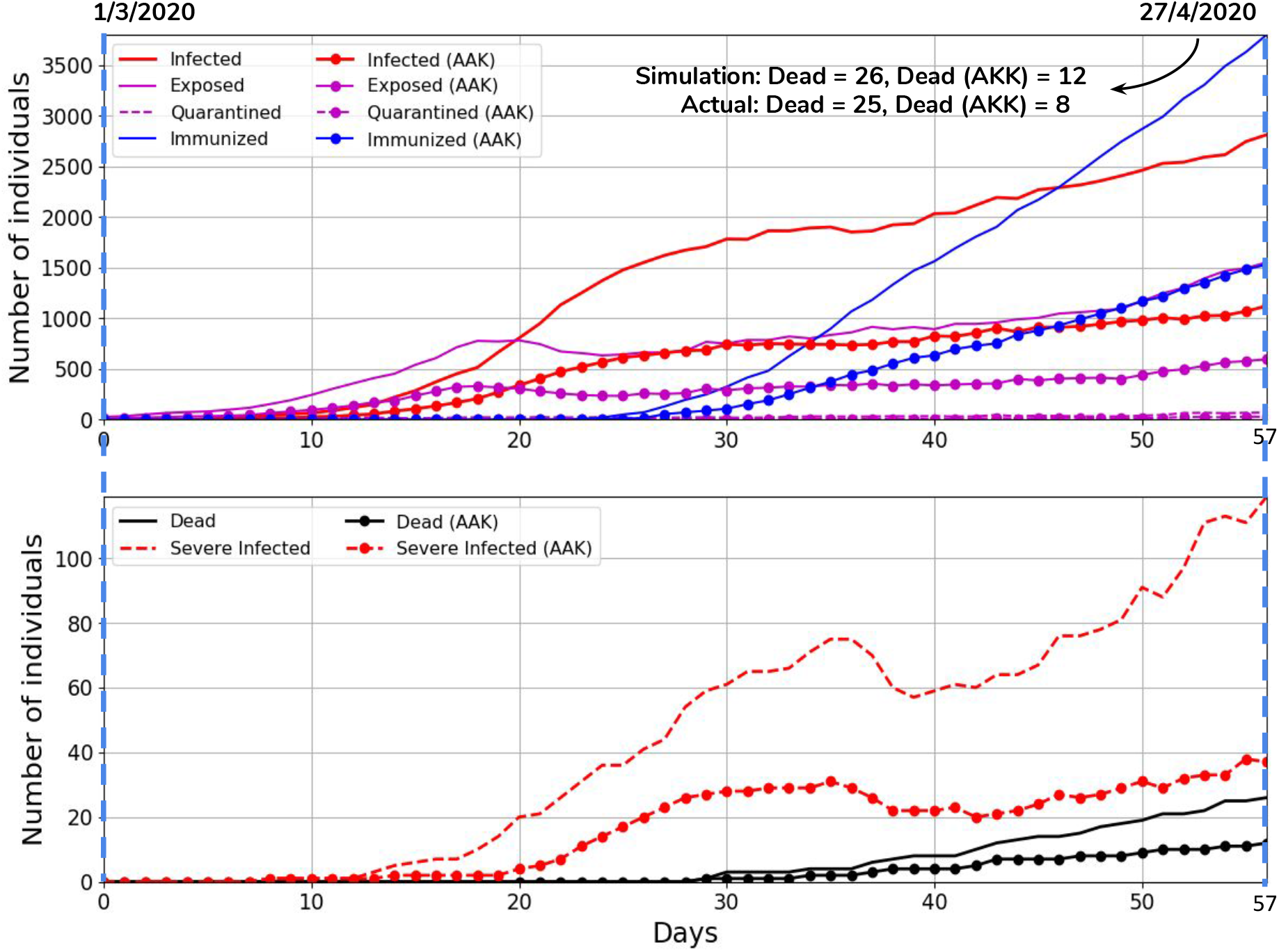
Simulation results for Almaty city and whole Kazakhstan from 1 March 2020 to 27 April 2020. Upper plot shows the number of Infected, Exposed, Quarantined and Immunized states versus time. The bottom plot shows the number deaths and severe infected versus time.

#### 2) COVID-19 Control Strategies

After establishing the initial conditions for Kazakhstan, and calibrating the state transitions, the next step was to model outcome scenarios for four variations of policy intervention: 1) complete quarantine, 2) going back to normal life, 3) “New Normal”, and 4) augmenting the “New Normal” strategy with the introduction of technological measures to detect infection zones and facilitate individual exposure detection and contact tracing.

The New Normal strategy introduces measures such as an increase in hospital capacity, unhindered supply of all necessary drugs, effective social distancing of vulnerable people, an increase in testing rates, and persistent observation of hygienic precautions and moderate social distancing by the general population.

The introduction of enhanced technological monitoring methods, such as identification of infection zones, and automated notifications to individuals whenever they have visited an infection zone or come into physical proximity with an infected individual, would increase the ability to detect and trace infections; in this scenario we used much higher quarantine rates due to presumed greater ability to control the interaction of individuals in the exposed and susceptible categories. The Fig. 6 shows the number of infected and dead versus time for these four strategies simulated from April 27. Table VIII summarizes the key parameters used in the simulations.

**Fig. 6:**
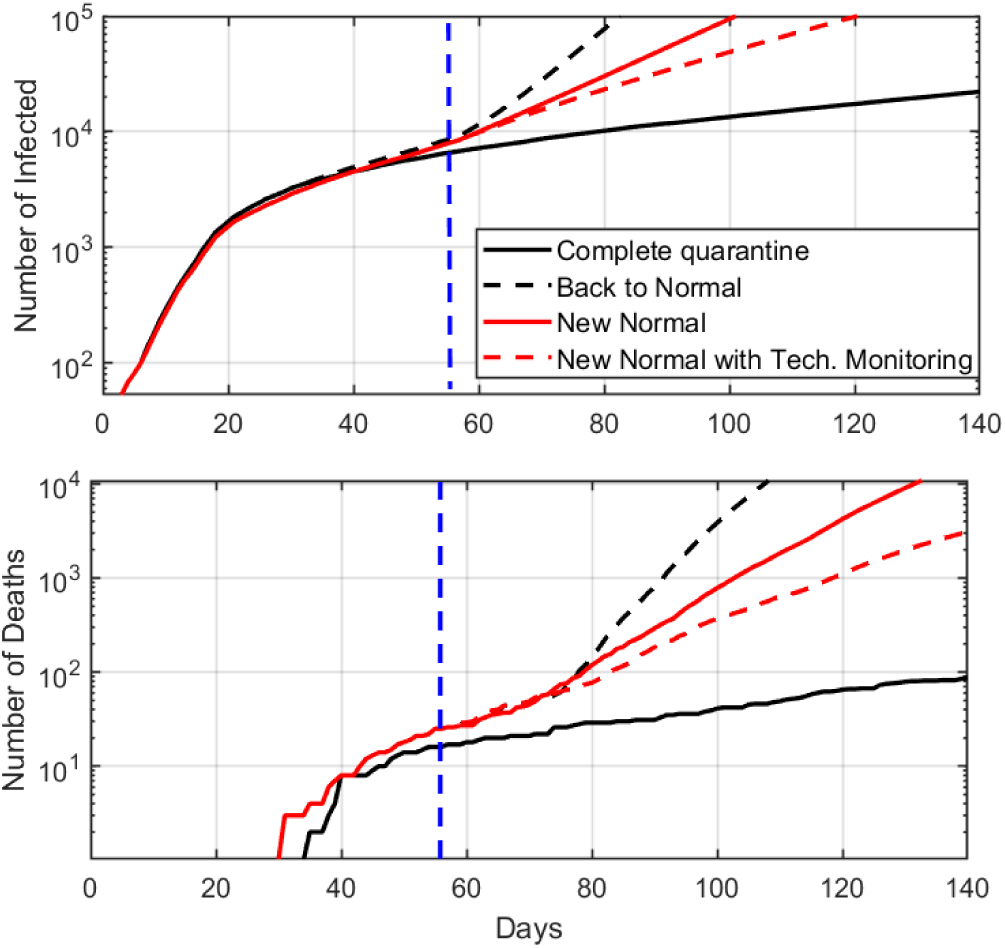
Simulation results showing infected and deaths versus time for the four control strategies.

**Table VIII:** Parameters for the Four Control Strategies

| Control Strategy | $\beta_{exp}$ | $tr$ | $qr$ |
| --- | --- | --- | --- |
| Complete Quarantine | 0.10 | 0 | 0.02 |
| Back to Normal | 0.16 | 1 | 0.02 |
| New Normal | 0.12 | 1 | 0.07 |
| New Normal with Technological Monitoring | 0.11 | 1 | 0.14 |

The results illustrate the effectiveness of the complete quarantine approach, and the impact of technological measures, compared to the New Normal strategy. Although the quarantine is demonstrably the most effective, it is not sustainable long-term due to extremely negative social and economic impact. As for the second strategy - back to normal lifestyle - simulation shows that it is also impractical due to an exponential second wave of epidemic growth in the country, thus effectively losing all benefits accrued from prior interventions. From these considerations the optimal policies would be a hybrid approach that balances the effectiveness of quarantine with measures to minimize the *b_exp_* value; the model could be used to establish this balance, ongoing, with adjustments made on a node-by-node basis.

#### 3) Intermittent Quarantine Strategy

The policies implemented in most countries including Kazakhstan are predominantly non-pharmaceutical suppression strategies designed to abruptly interrupt viral propagation (temporarily reducing *R*_0_), and thereby “flatten the curve” to minimize impact on the healthcare infrastructure until such time as pharmaceutical or immunological outcomes can be achieved.

The suppression strategies consist of strict quarantine measures such as the lockdown of specific regions, banning of social gatherings, closure of schools and universities (and subsequent shift to fully on-line delivery methods), national adoption of work-from-home policies, and the temporary closure of non-essential productions.

To achieve the aforementioned balance, we considered an intermittent quarantine regime, effectively alternating between suppression and controlled mitigation strategies with β*_exp_* values of 0.19 and 0.05 respectively for quarantine off and on periods (see Fig. 7).

**Fig. 7:**
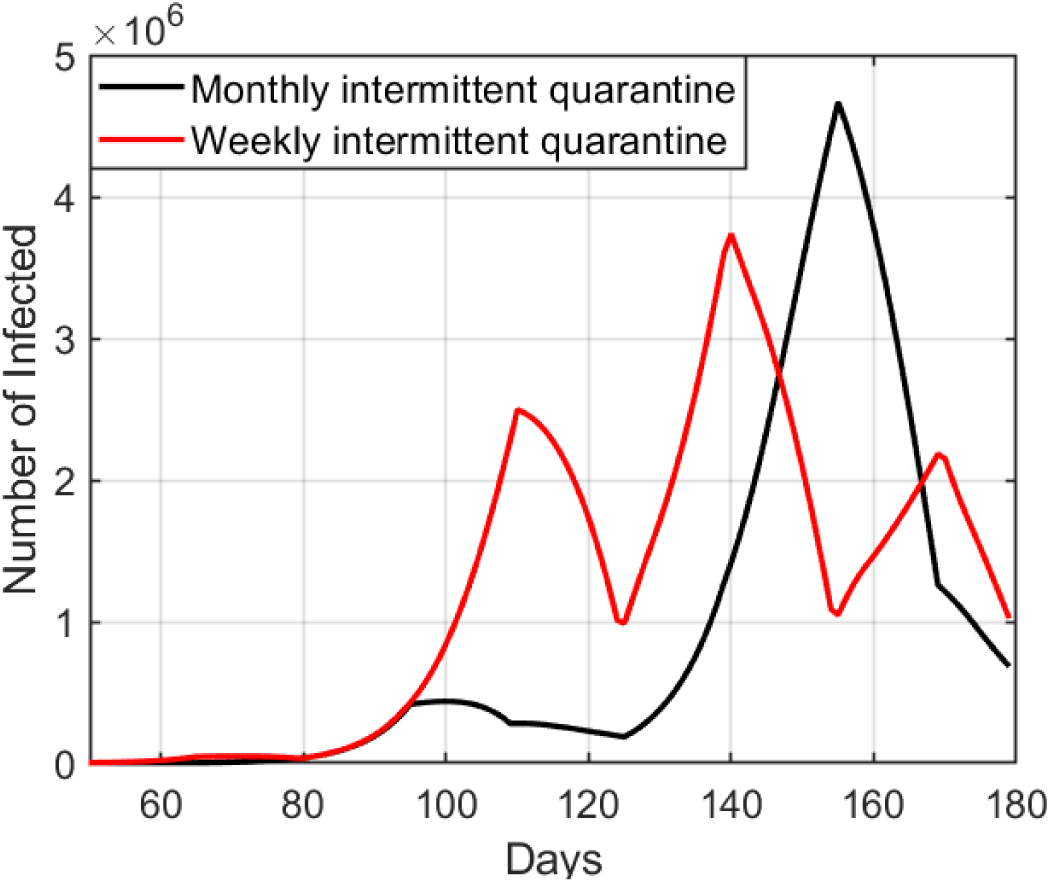
Number of infected versus time for intermittent quarantine strategy simulations with monthly and weekly intervals.

The simulation analysis shows that for longer periods of intermittent quarantine the peak in infected is much higher but the number of infected (area under the curve) is lower; the analysis is conducted as a multi-objective optimization problem as a function of time and medical preparedness.

#### 4) Effects of Transportation Limitations

Our simulator incorporates domestic population mobility based on the transition matrix. The inclusion of mobility makes the model more realistic, with more nuanced modeling of suppression policy effects, and thus the simulation results more precise. To illustrate the effect of transportation limitations on the infection spread, we simulated the validation scenario with four different traffic ratios (0, 0.33, 0.66 and 1) starting from day 57. The peak of infected numbers increases with traffic ratio as shown in the Fig. 8.

**Fig. 8:**
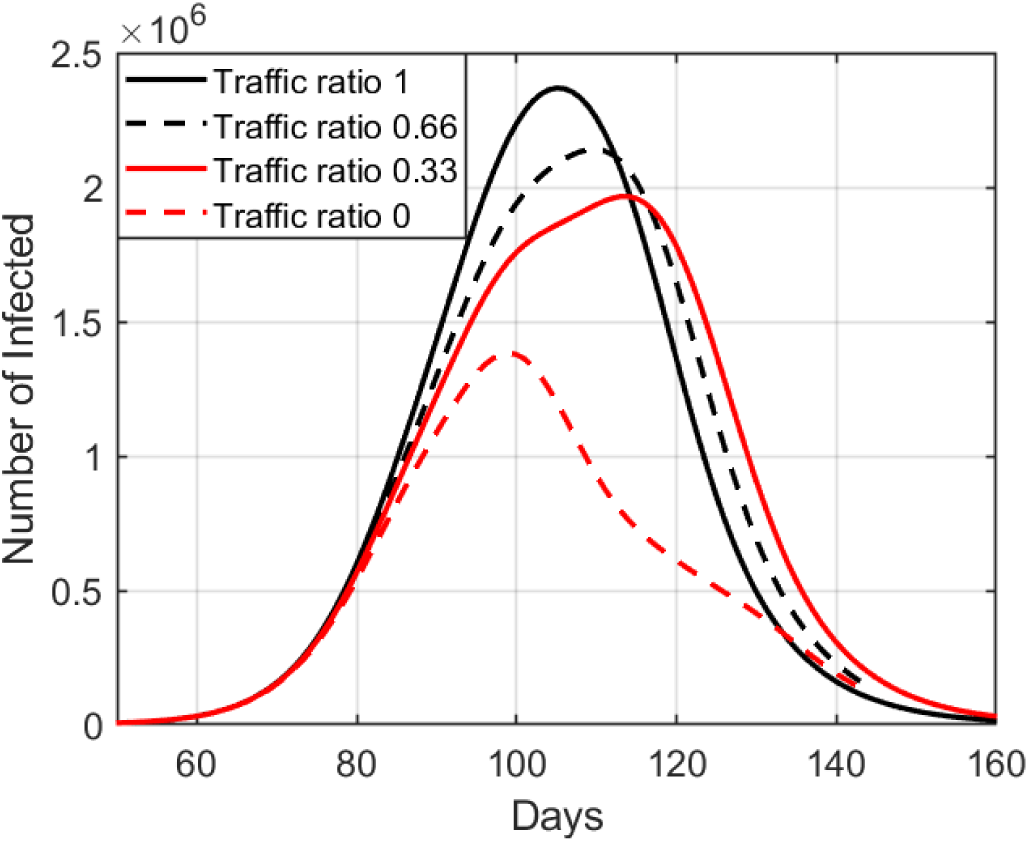
Effect of transportation limitations on the epidemic spread.

The traffic ratio of zero illustrates the case of complete quarantine, ongoing, which might not be feasible for both practical and economic reasons.

## V. Discussion

The network-of-nodes design of the stochastic epidemic simulator allows granular adjustments to the model’s parameters at the level of individual regions (nodes), and the interactive controls allow for testing policy interventions at discrete time intervals, also, unique to each region. The inclusion of mobility transitions between regions also allow for more precise simulation of events.

The calibration of the model presents a conundrum, in that if we fit the parameters of the model to the reported data on confirmed cases, and set the state transition rates based on those reports, then the simulator outcomes will be dependent on the relative accuracy of the reported data, and factors such as physical limits on the number of tests that can be conducted could skew the outcomes.

In the case of COVID-19, it is likely that the number of infected individuals, and the number of deaths attributed to it, are higher than the number of laboratory-confirmed cases, given the relatively uneven rates of testing across different populations, and wide variations in percentage of positive tests, along with the variations in mortality rates, and the concurrent excess mortality observed in many of the reporting zones. Perhaps the single greatest factor is that most countries only test if a patient meets screening criteria, with explicit symptoms, but exposed patients can remain asymptomatic for approximately five days, and a non-trivial percentage of those infected may not seek medical care, due to light symptoms, and the infection may not be recognized or registered as COVID-19 due to sensitivity and specificity of rapid tests available on market [36].

We have run many scenarios, and, often, the projections of the simulator ran higher than the reported figures. This was an initial concern, until updated reporting inclusive of testing rates, positive rates, mortality and excess mortality were taken into account, which suggested that the preliminary reports were simply incomplete. Thus, if we run the simulator tightly, calibrating and updating based only on the reported confirmed case data, then we risk essentially modeling for those with severe enough symptoms that they require medical care, but by doing so the model will only capture a portion of the impact, and skew the population sizes for the Susceptible and Recovered states. The significance of this outcome becomes greater over time, as it determines whether and when a population reaches herd-immunity levels, and so too, the duration and extent of NPI policies.

In order to overcome this outcome it is necessary to establish a more rigorous testing regimen, in accordance with standard testing protocols, to accurately estimate the size of each population state.

In this case the advantage of the node-based approach lies in the opportunity to adjust the policies at the regional level, and thereby reduce the potentially unnecessary economic and social disruption of a country-wide lockdown, and strategically smooth the pressure on hospitals in different regions.

## VI. Conclusions

We have designed and implemented an open-source network-based stochastic epidemic simulator that models the movement of a disease through the SEIR states of a populace. The simulator incorporates interactive controls of state transition probabilities and modification of environmental factors such as health care capacity and mobility amongst regions.

The interactive controls enable the dynamic introduction of events and the testing of policy options that allow the evaluation of alternative scenarios and projection of their potential impact over time, at both the national and regional levels.

The simulator was calibrated with recent data on COVID-19 in the Lombardy region of Italy, utilized to fine-tune model assumptions regarding viral propagation and mortality rates in both normal and health-care over-capacity situations.

The simulator was then configured to model the COVID-19 outbreak in Kazakhstan, using empirical observations and authoritative data on initial conditions as provided by government sources, as part of a collaboration established with the express objective to inform policy deliberations.

Based on this Republic of Kazakhstan configuration, we successfully modeled the spread of the disease, and the impact of various stages of government policy decisions, such as closing schools, selectively quarantining regions, and the imposition of “lockdown” conditions (also known as “shelter in place”).

The simulator was then used to evaluate a range of scenarios with the goal of minimizing strain on the health care system while reducing negative social and economic impact by seeking a balance between containment of disease spread and the imposition of less stringent social controls on a localized basis.

On one extreme, the simulator indicates that a lack of mitigation will yield unmitigated disaster, with overwhelmed health care systems and high rates of mortality. On the other extreme, persistent suppression (via quarantines and strict social distancing) will curtail disease propagation and reduce mortality, but also cause economic harm, and instigate other forms of social distress.

The projections of the simulator suggest that going forward, it will be necessary to maintain some level of social controls guided by a comprehensive testing regimen so as to constrain propagation while minimizing social and economic impact, until such time as more widespread immunity amongst the population (so-called “herd” immunity) is gradually achieved, or a vaccine is introduced.

## Data Availability

The source code of our simulator was uploaded to GitHub under the BSD license.

https://github.com/IS2AI/COVID-19-Simulator

## Footnotes

1 *https://www.youtube.com/channel/UCr7o_0wW4nkqx-G5b7Zopgw*

2 github.com/IS2AI/COVID-19-Simulator

